# Validation of self-collected buccal swab and saliva as a diagnostic tool for COVID-19

**DOI:** 10.1101/2020.10.03.20205278

**Authors:** Chee Wai Ku, Durai Shivani, Jacqueline Q T Kwan, See Ling Loy, Christina Erwin, Karrie K K Ko, Xiang Wen Ng, Lynette Oon, Koh Cheng Thoon, Shirin Kalimuddin, Jerry KY Chan

## Abstract

**Background:** Effective management of Severe Acute Respiratory Syndrome Coronavirus 2 (SARS-CoV-2) requires large-scale testing. Collection of nasopharyngeal swab (NPS) by healthcare workers (HCW) is currently used to diagnose SARS-CoV-2, which increases the risk of transmission to HCWs. Self-administered saliva and buccal swabs are convenient, painless and safe alternative sample collection methods.

**Methods:** A cross-sectional single centre study was conducted on 42 participants who were tested positive for SARS-CoV-2 via NPS within the past 7 days. A self-collected saliva and buccal swab and a HCW-collected NPS were obtained. Real-time polymerase chain reaction (RT-PCR) was performed and cycle threshold (CT) values were obtained. Positive percent agreement (PPA), negative percent agreement (NPA) and overall agreement (OA) were calculated for saliva and buccal swabs, as compared with NPS.

**Results:** Among the 42 participants, 73.8% (31/42) tested positive via any one of the 3 tests. With reference to NPS, the saliva test had PPA 66.7%, NPA 91.7% and OA 69.0%. The buccal swab had PPA 56.7%, NPA 100% and OA 73.8%. Presence of symptoms improved diagnostic accuracy. There was no statistically significant association between CT values and duration of symptom onset within the first 12 days of symptoms for all three modalities.

**Conclusion:** Self-collected saliva tests and buccal swabs have only moderate agreement with HCW-collected NPS swabs. Primary screening for SARS-CoV-2 may be performed with a saliva test or buccal swab, with a negative test warranting a confirmatory NPS to avoid false negatives. This combined strategy minimizes discomfort and reduces the risk of spread to the community and HCWs.

## BACKGROUND

Severe Acute Respiratory Syndrome Coronavirus 2 (SARS-CoV-2), the causative agent of Coronavirus Disease 2019 (COVID-19), was first identified in December 2019 [1]. In March 2020, COVID-19 was declared a pandemic by the World Health Organisation.

One of the main reasons for the high rate of transmission is the significant proportion of asymptomatic but infective carriers. Hence safe and effective detection of asymptomatic COVID-19 patients through appropriate large-scale testing is of paramount importance.

As the clinical symptoms COVID-19 patients experience are often nonspecific, the current method of detection relies heavily on molecular techniques. It is recommended that samples for testing are obtained from the upper respiratory tract rather than lower respiratory tract [2]. These include nasopharyngeal swabs (NPS), the current standard test, as well as oropharyngeal swabs, saliva specimen, and nasal aspirates.

The accuracy of COVID-19 detection varies according to the viral load in the different respiratory tract samples. In the first 14 days after onset of illness, SARS-CoV-2 was most reliably detected in sputum samples, which contained the highest viral load, followed by nasal swabs [3, 4]. There are several disadvantages of the NPS, which is considered the current standard test “gold-standard” test in most settings. Firstly, NPS can only be performed by trained HCWs. The patient needs to travel to the swabbing facility which increases the risk of community spread. Secondly, to perform the swab, the medical staff must be in close contact with the patient. Coughing or retching by the patient could produce a large number of aerosolised droplets, increasing the risk of HCW transmission [5]. Thirdly, this increases the burden on the currently heavily strained healthcare system by diverting a lot of resources to the diagnosis of SARS-CoV-2. Furthermore, a significant proportion of suspect cases who reside in the community are asymptomatic and are only called up for testing as a result of contact tracing from a confirmed COVID-19 case and hence are unlikely to present for testing. Hence, our group seeks to validate diagnostic tests which can be performed by the patient at home, and if validated, may have comparable concordance to NPS.

In instances where an NPS is not possible, the Centers for Disease Control and Prevention (CDC) and the Food & Drug Administration (FDA) recommend a HCW-collected oropharyngeal specimen as other alternatives. Indeed, active viral replication in the upper respiratory tract of young to middle-aged patients with mild cases of COVID-19 has been demonstrated previously, with peak viral shedding during the first week of symptoms [6]. There is also emerging evidence to suggest that supervised self-collected oral fluid specimens perform similarly to HCW-collected NPS specimens for the detection of SARS-CoV-2 infection [7].

In order to validate the use of buccal swabs and saliva specimen as alternative diagnostic tests for SARS-CoV-2, our group performed a cross-sectional study of NPS, self-collected buccal swabs and saliva specimens collected concurrently in order to determine the positive percent agreement (PPA), negative percent agreement (NPA), overall agreement (OA), positive and negative predictive values. The advantage of self-collected buccal swabs or saliva specimen is two-fold, 1) facilitates specimen collection without patients leaving their home, thus improving compliance and ease of collection, and 2) reduces the risk of community spread and transmission to HCWs. This would revolutionize the management of SARS-CoV-2, where suspect cases can send in a specimen for testing without breaching quarantine notice, and thus increase detection rates without compromising the safety of others.

## METHODS

### Setting and participants

A cross-sectional single centre study was conducted on 42 individuals who were previously tested positive for SARS-CoV-2 via NPS within the past 7 days, and who were isolated at the Singapore General Hospital (SGH). SGH is the largest tertiary hospital in Singapore and is one of the main referral hospitals for treating COVID-19 patients.

The inclusion criteria were participants diagnosed with SARS-CoV-2 infection by NPS, between the ages of 21-80 years. Patients who were unable to produce oral secretions for self-collection were excluded from the study. Written informed consent was obtained. This study was approved by the institution’s ethics review board (CIRB Ref No 2020/2655).

### Demographic, clinical data collection and survey on patient experience

Sociodemographic data and symptoms at time of sampling were obtained via a questionnaire administered by a study team member, and through review of medical records. After sample collection was complete, the patient’s experience was surveyed. They were asked to select the test that best fit each of the following qualities: 1) comfort 2) convenience and 3) personal preference.

### Sample collection and processing

A saliva, buccal and NPS sample was obtained from each participant in that order. For the saliva specimen, participants were asked to cough deeply five times and pool saliva in their mouth for 1-2 minutes prior to collection, and gently spit 1-2 mL of saliva into a 60 mL sterile closed-top plastic collection container (BMH.921406, Biomedia, Singapore). Subsequently, for the buccal sample, participants were asked to pool any phlegm or secretions in their mouth, rub the swab (300264, Deltalab, Spain) on both cheeks, above and below the tongue, both gums, and on the hard palate for a total of 20 seconds to ensure the swab was saturated with oral fluid. The swab was then placed in the tube with the lid secured. Finally, the NPS (MSC-96000-ST, Miraclean, China) was collected by a trained HCW for all patients, as per standard hospital protocol [8]. All swabs were processed in 1ml of lysis buffer (Cobas Omni Lysis Reagent, P/N 06997538190) and in-house RT-PCR was performed on all specimens based on protocol by Corman et al [9]. The results for SARS-CoV-2, including the E-gene cycle threshold (CT) values, were correlated to that for the NPS. All signals that crossed the detection threshold were considered a positive test.

### Statistical analysis

Independent t-test or Mann-Whitney test was applied to continuous variables as appropriate, and Fisher’s exact test was applied to categorical variables. Results of the saliva and buccal swabs were individually compared to the NPS and the positive percentage agreement (PPA), negative percentage agreement (NPA), overall agreement (OA), positive predictive values (PPV) and negative predictive values (NPV) were calculated. CT values between the saliva and buccal swabs with NPS were analysed with the paired t-test. Statistical analyses were conducted using SPSS Statistics Version 20.0 (IBM Corp, Armonk, NY, USA).

### RESULTS

All the patients who met the inclusion criteria were recruited. Among 42 participants who previously tested positive for SARS-CoV-2 via a nasopharyngeal swab (NPS) within the preceding 7 days, 73.8% (31/42) tested positive via any one of the 3 swab tests. Table 1 shows the baseline demographics between those who tested positive or negative, for the three diagnostic tests for SARS-CoV-2, namely NPS, buccal and saliva (Table 1). Participants who remained NPS positive at the time of study recruitment were younger, and more likely to be symptomatic (Table 1). The survey of participant experience showed that 59.5% (25/42) participants ranked the buccal swab as the most preferred, most convenient, as well as most comfortable to collect. A total of 40.5% (17/42) participants chose the saliva test as the most preferred, convenient, and comfortable means for first-line SARS-CoV-2 testing. Only one participant ranked NPS as most preferable (1/42), while none of the them felt that it was convenient or comfortable.

**Table 1.**
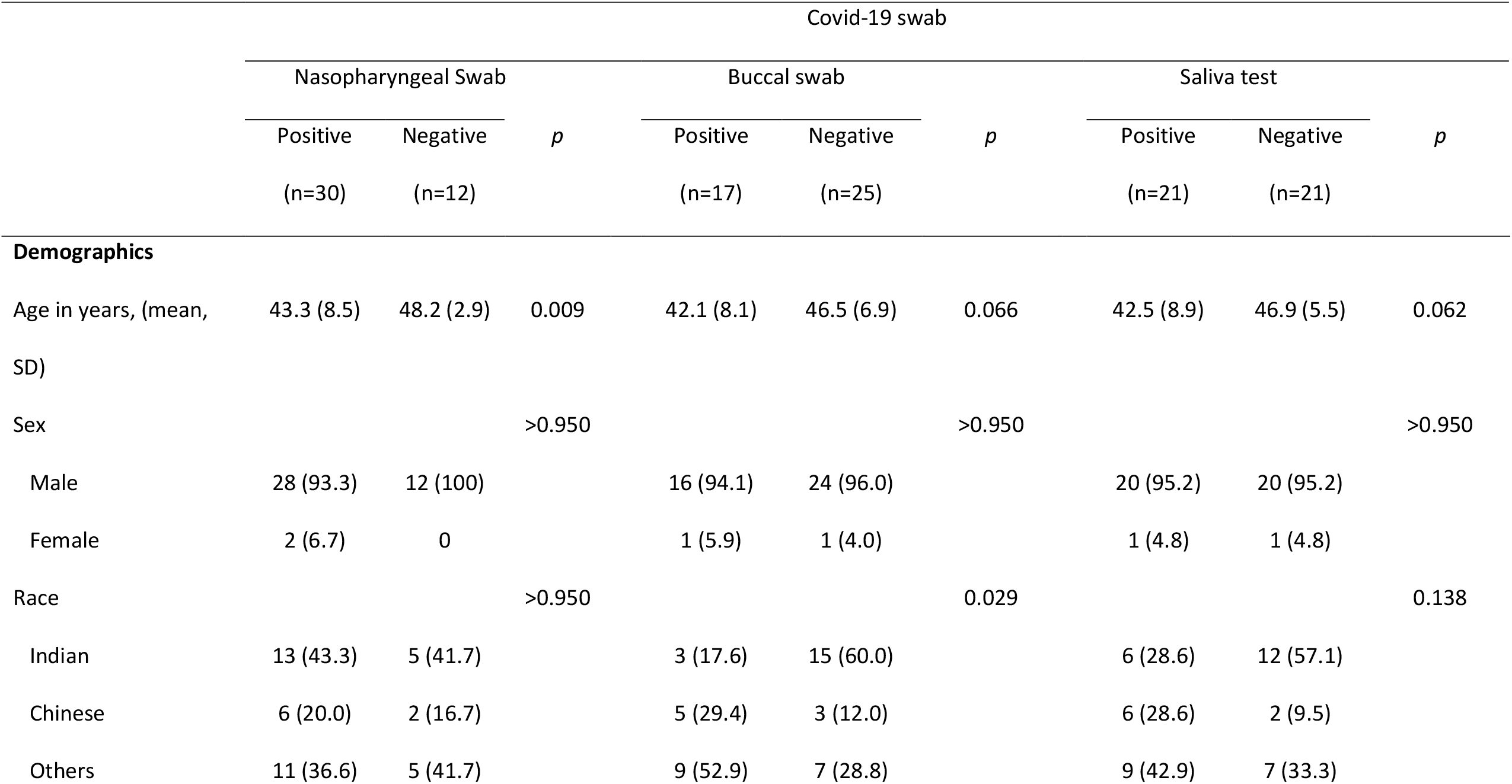
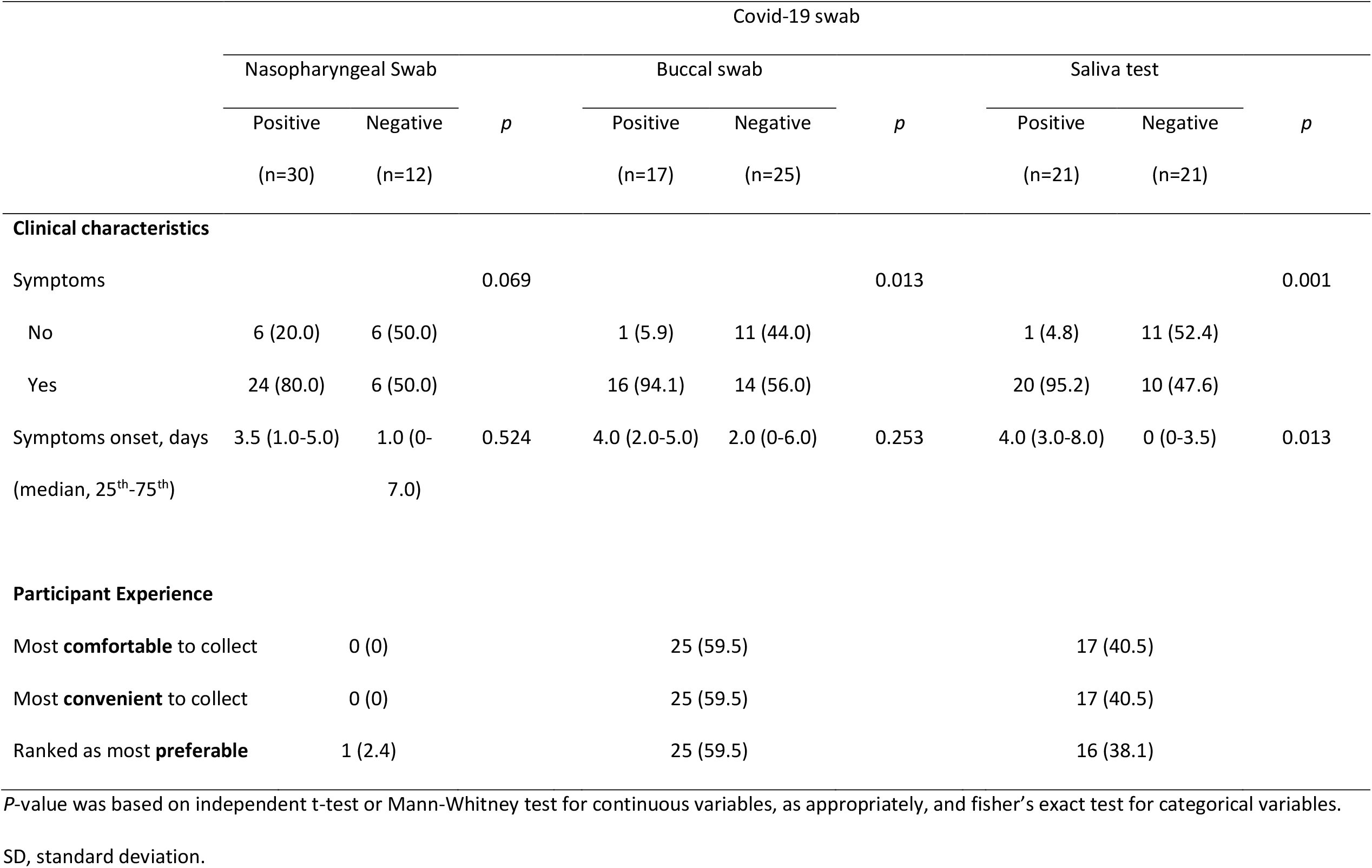
Baseline demographics, clinical characteristics of patients and the results of paired nasopharyngeal, buccal and saliva test and patient experience survey performed (n=42) *P*-value was based on independent t-test or Mann-Whitney test for continuous variables, as appropriately, and fisher’s exact test for categorical variables.

The incidence of SARS-CoV-2 in this cohort as diagnosed using NPS was 71.4% (n=30/42). With reference to the NPS, the buccal swab had a PPA of 56.7%, NPA of 100%, OA of 73.8%, PPV of 100% and NPV of 48% (Table 2A). Viral load was lower in the buccal specimen, with the mean CT value for the buccal swab being higher than NPS (27.19 ± 2.48 vs 21.66 ± 5.60, p < 0.001) (Figure 1A). With reference to the NPS, the saliva sample had a PPA of 66.7%, NPA of 91.7%, OA of 69.0%, PPV of 95.2% and NPV of 52.4% (Table 2B). There was no difference in CT values between saliva and NPS specimens (25.77 ± 5.60 vs 22.95 ± 6.03, p = 0.057), suggesting a similar viral load in both samples (Figure 1B).

**Table 2.** Comparison between nasopharyngeal and buccal swab (a), and nasopharyngeal and saliva test (b) with positive percent agreement, negative percent agreement, positive predictive value and negative predictive value.

| Nasopharyngeal swab |  |  |  |  |
| --- | --- | --- | --- | --- |
|  |  | Positive | Negative |  |
| (a) Buccal swab | Positive | 17 | 0 | Positive predictive value = 100% |
|  | Negative | 13 | 12 | Negative predictive value = 48.0% |
|  |  | Positive Percent Agreement = 56.7% | Negative Percent Agreement = 100% | Overall Agreement = 69.0% |
| Nasopharyngeal swab |  |  |  |  |
|  |  | Positive | Negative |  |
| (b) Saliva test | Positive | 20 | 1 | Positive predictive value = 95.2% |
|  | Negative | 10 | 11 | Negative predictive value = 52.4% |
|  |  | Positive Percent Agreement = 66.67% | Negative Percent Agreement = 91.7% | Overall Agreement = 73.8% |

**Figure 1.**
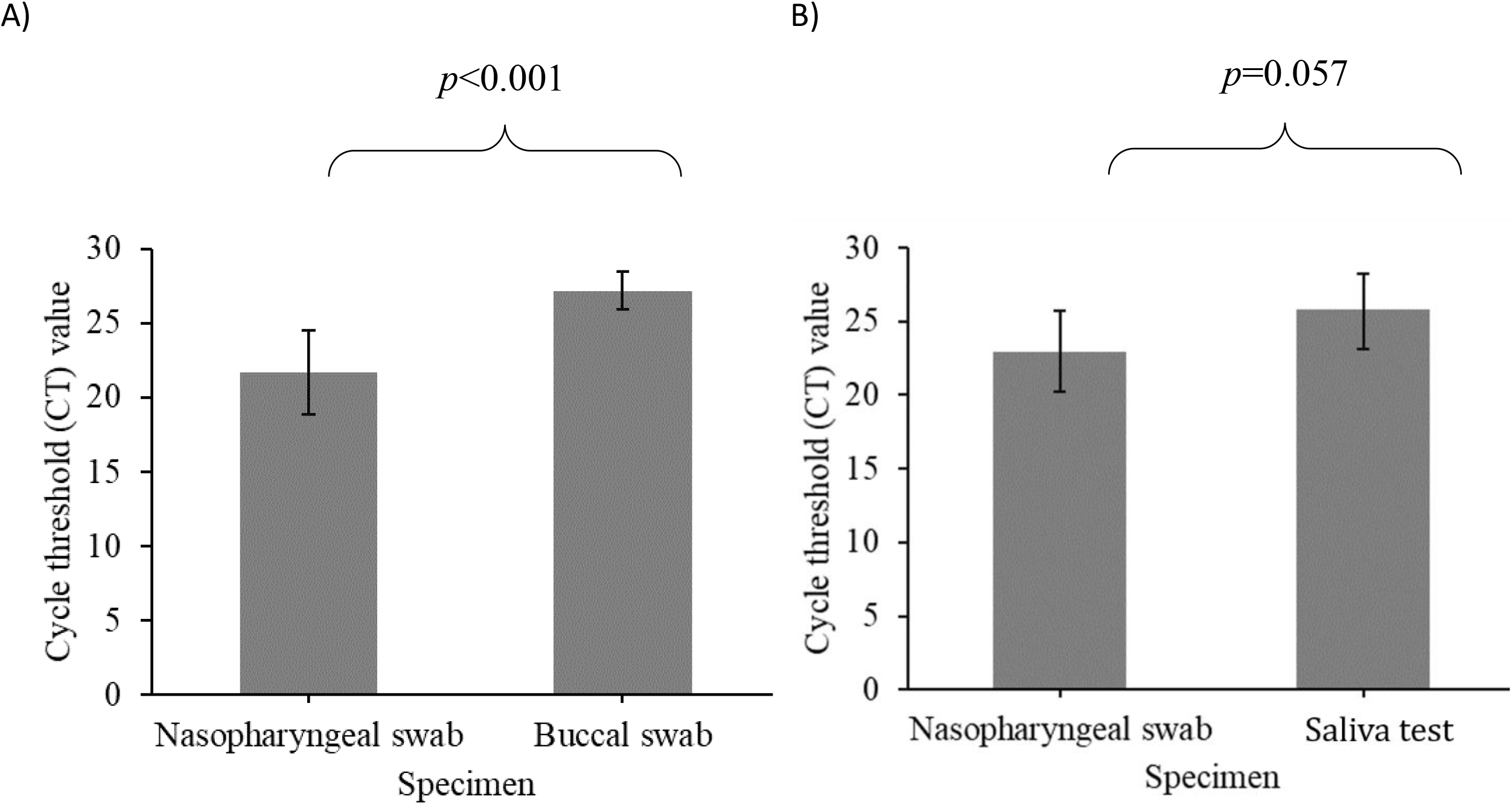
Comparison of cycle threshold values between nasopharyngeal swab and buccal swab (A) and between nasopharyngeal swab and saliva test (B). Error bars represent 95% confidence intervals of the mean values. P-value was based on paired-t test.

Presence of symptoms at the time of swab collection was associated with better diagnostic accuracy. 53.3% of symptomatic participants tested positive by both NPS and buccal swab (p = 0.017), while 63.3% were positive by both NPS and saliva sample (p = 0.004) (Table 3). There was no statistically significant association between CT values and duration of symptom onset, for all three diagnostic modalities (Figure 2). Comparison of CT values for the paired NP, saliva and buccal swabs are shown in Figure 3.

**Table 3.**
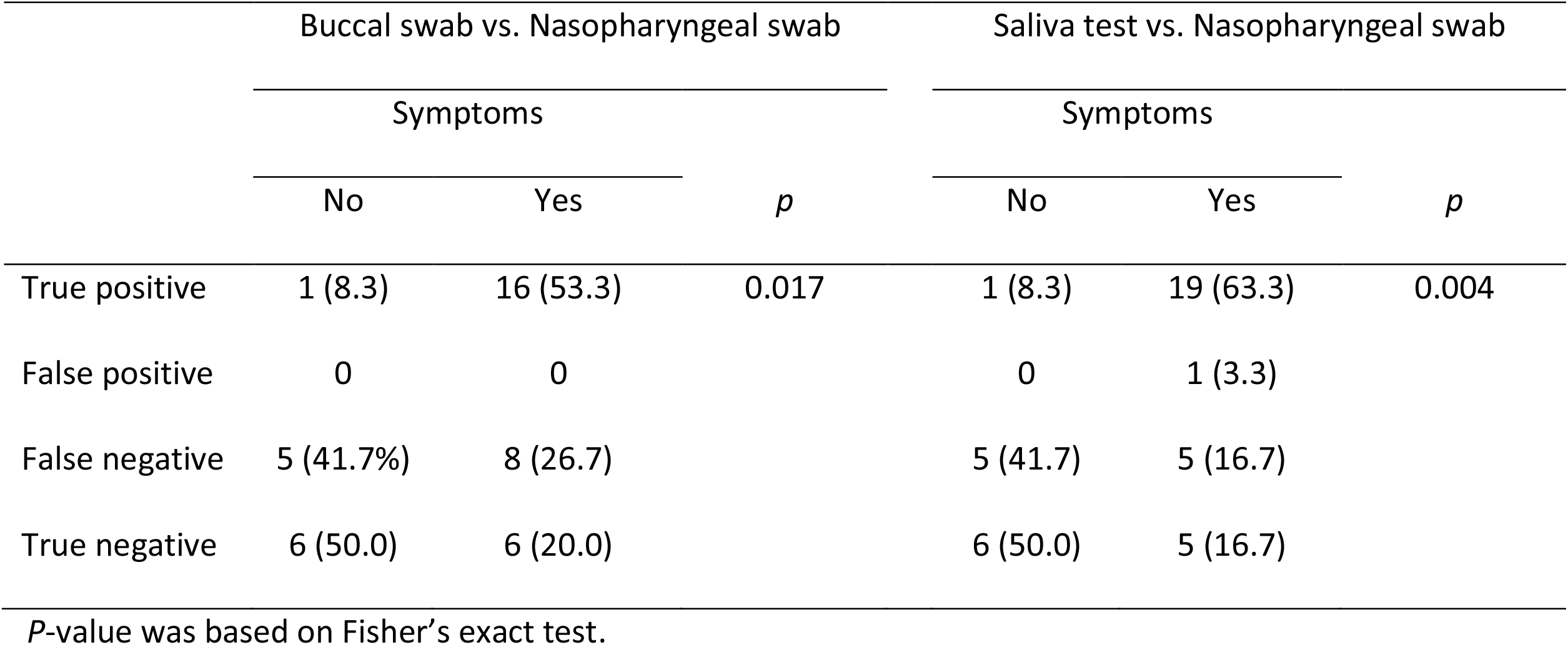
The associations between diagnostic tests and presence of symptoms

|  | Buccal swab vs. Nasopharyngeal swab |  |  | Saliva test vs. Nasopharyngeal swab |  |  |
| --- | --- | --- | --- | --- | --- | --- |
|  | Symptoms |  | *p* | Symptoms |  | *p* |
|  | No | Yes |  | No | Yes |  |
| True positive | 1 (8.3) | 16 (53.3) | 0.017 | 1 (8.3) | 19 (63.3) | 0.004 |
| False positive | 0 | 0 |  | 0 | 1 (3.3) |  |
| False negative | 5 (41.7%) | 8 (26.7) |  | 5 (41.7) | 5 (16.7) |  |
| True negative | 6 (50.0) | 6 (20.0) |  | 6 (50.0) | 5 (16.7) |  |

*P*-value was based on Fisher's exact test.

**Figure 2.**
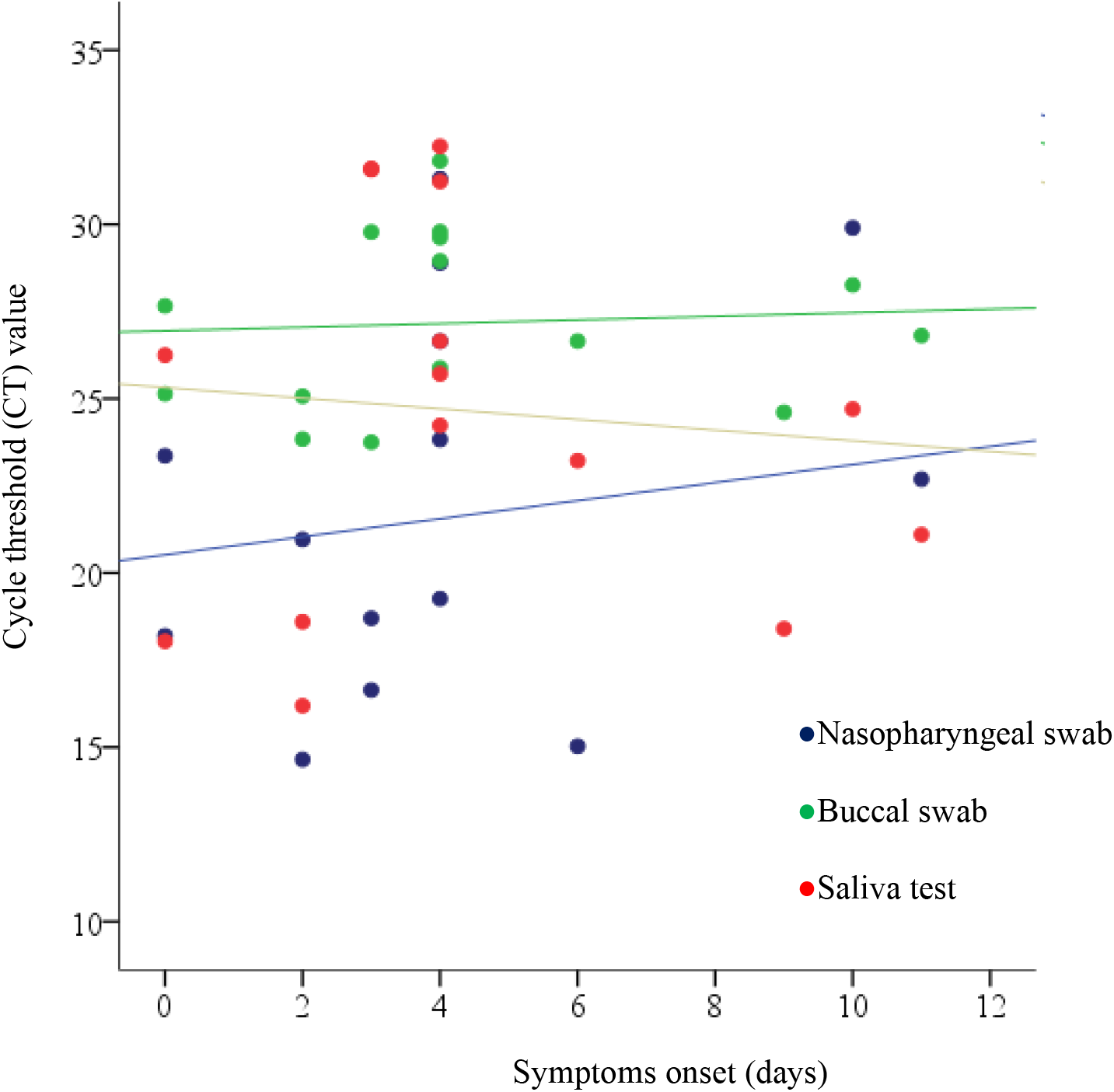
The associations between cycle threshold values of nasopharyngeal, buccal and saliva tests with duration of symptom onset. CT of nasopharyngeal swab and days: r=0.14 (p=0.485); CT of buccal swab and days: r=0.07 (p=0.806); CT of saliva test and days: r=0.171 (p=0.471).

**Figure 3.**
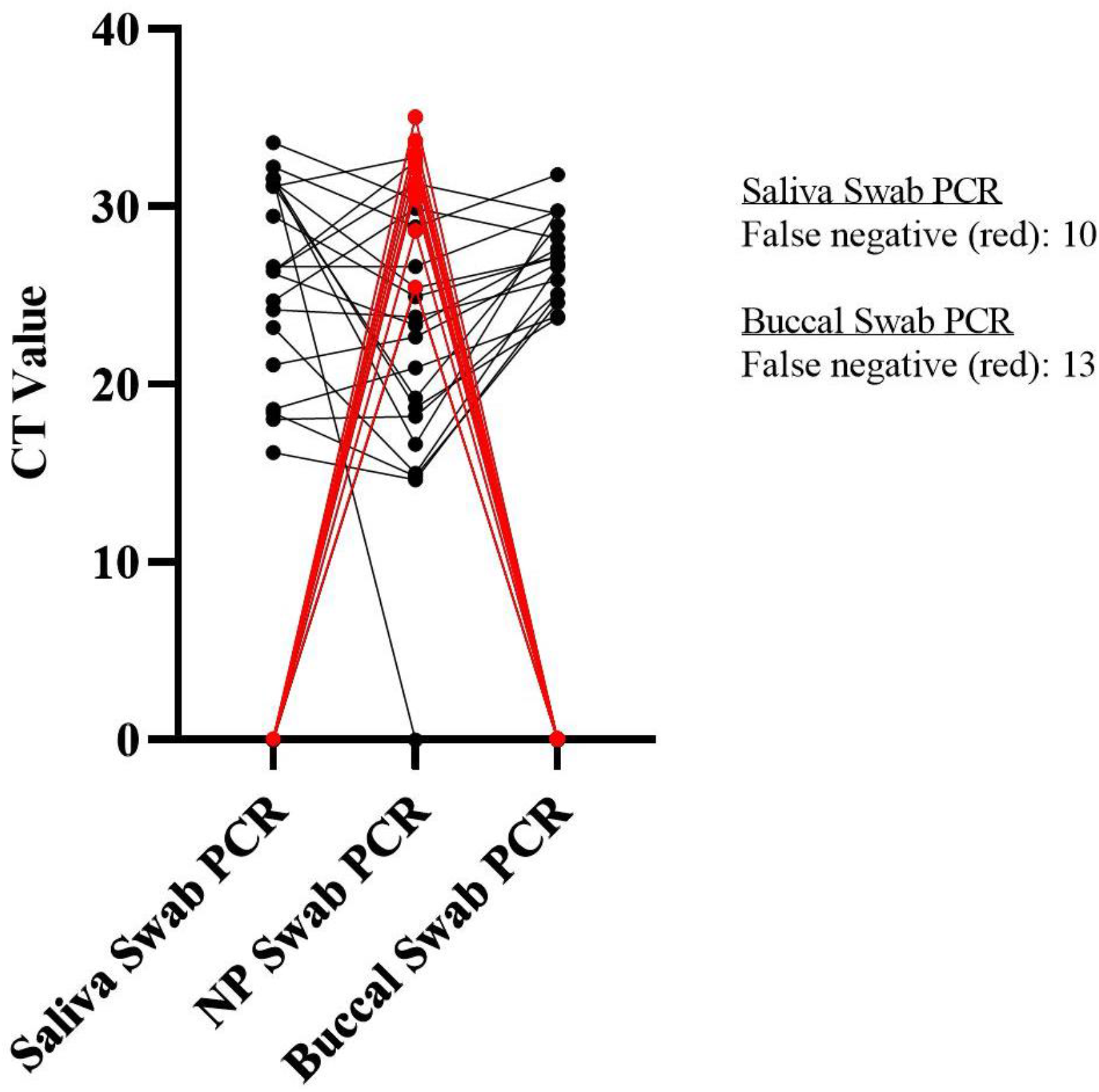
Comparison of CT value for paired nasopharyngeal, saliva and buccal swabs performed in SARS-CoV-2 positive patients

## DISCUSSION

In this study, we have shown that saliva tests and buccal swabs were comparable to each other and were in moderate agreement with NPS for the detection of SARS-CoV-2, with PPA between 56 and 66% and PPV 95 to 100%. On the other hand, both saliva tests and buccal swabs performed comparably to NPS in detecting negative cases, with an NPA of 90 to 100%. Overall, they were moderately comparable to NPS with an OA of 69 to 74%. This paves the way for larger validation studies to support the use of self-collected saliva tests or buccal swabs, without the risk of community spread or spread to HCW.

Most coronaviruses are known to replicate in the epithelial cells of the respiratory tract. The viral shedding pattern in SARS-CoV-2 has been shown to be similar to that of the influenza virus, with peak viral shedding in the first week of illness [6, 10]. NPS has been shown to have better diagnostic accuracy compared to oropharyngeal swab [10-12], and it has been recommended as the gold standard diagnostic modality. However, there is emerging evidence alluding to similar detection rates with alternative sample collection methods that may be feasible and safe, including saliva, posterior oropharyngeal saliva, and throat washing.

Self-collected saliva offers a promising prospect for sample collection. This is due to its relative convenience, comfort and subjective participant preference compared to the NPS. It is also less costly and quicker to obtain [13], with the additional benefit of decreased risk of transmission to HCWs, making it a convenient and safe means of mass testing. There is increasing evidence that the viral load in saliva is comparable or higher than that in the nasopharynx [4, 14-16]. We have shown that the CT values were comparable between saliva and NPS, representing a similar viral load in both samples. A longitudinal study in Korea showed that the highest viral load was in the nasopharynx, but also remarkably high in the saliva, and was detected in the saliva up to day 6 of hospitalization and day 9 of illness [4]. Azzi et al reported a high overall concordance rate of 97.4% between NPS and saliva, and also showed that there were no statistically significant difference between viral loads of the two samples [15, 17]. Our results also showed a high PPV of 95.2% compared to NPS, the current standard diagnostic test for SARS-CoV-2. However, the low PPA of 66.7% may limit its suitability to replace NPS as the gold standard diagnostic test. Existing studies have also shown varying PPA or concordance rates, when comparing saliva detection rates to NPS, with reported positive concordance rates ranging from 45.6% [14] to 94.8% [18]. The vast differences in concordance rates may be due to limitations in sample collection. The lower PPA might also be limited by the amount and sample collection technique of the saliva test. Moreover, considering the similar viral load between saliva and NPS, this further supports that poor sample collection technique might be the reason leading to some saliva tests being completely negative. Unfortunately, our study did not investigate the quality of the saliva produced. Kojima et al observed that there were differences in positive detection rates between clinician supervised vs unsupervised self-collected saliva tests and NPS, which further supports that collection technique plays a big role in the positive detection rates [7]. Another possible explanation is the duration from diagnosis to sample collection. Wyllie et al has shown that a higher percentage of saliva samples remained positive up to 10 days after the COVID-19 diagnosis compared to NPS (81% vs 71%)[19], which is a result supported by other studies [20, 21]. Although our results show a non-significant relationship between interval after symptom onset and sample collection, other studies have shown a possibility that NP and saliva are most equivalent early in the illness compared to those collected beyond the first week [22]. Not surprisingly, the presence of symptoms increased the PPA for saliva compared to NPS (79.2%, 19/24). Overall, these findings provide supporting evidence for recommending saliva as an alternative modality that is safer, more comfortable and convenient to collect, and sufficiently accurate for making a clinical diagnosis.

There are very few studies evaluating the use of buccal swabs, with conflicting results between studies [7, 22, 23]. Our results show that the buccal swab performs less accurately than NPS due to its lower PPA rate of 56.7%. It also has a lower viral load as reflected in the significantly higher CT value obtained. However, participants in our study ranked it as the most convenient, comfortable and preferred test.

Thus, buccal swabs may have a role in specific populations who might not be able to spontaneously produce saliva, such as in young children and older patients [23].

The correlation of detection rates and symptoms still remains controversial. Zou et al showed that viral load in asymptomatic patients was similar to symptomatic patients with NPS and throat swabs [10], while Chau et al showed that viral loads were equivalent in symptomatic patients, but lower in asymptomatic patients in saliva [24]. We showed that none of the tests showed evident association with duration of symptom onset. However, symptomatic patients were more likely to be true positive on both the buccal swab and saliva test.

Taken together, buccal and saliva samples have moderate agreement with NPS, and are reasonable alternatives to the current gold standard NPS for diagnosis of COVID-19. In view of the comparable viral load, high PPV and OA, moderate PPA, and greater patient comfort and convenience, we recommend that the initial screening NPS be replaced with a saliva test or buccal swab for community testing. If the result is positive, we can assume that the patient is truly SARS-CoV-2 positive, due to their high PPV when compared to NPS. If negative, we should perform a confirmatory NPS before discharging the patient, to mitigate the moderate PPA value to ensure they are truly COVID-negative (Figure 4). This workflow is time and resource-saving, especially in the context of mass screening strategies where resources, in particular HCWs, are scarce. In this context, saliva tests or buccal swabs can easily increase testing rates due to their ease of collection, without further straining the healthcare system. The requirement for a confirmatory NPS for negative cases combines the sensitivity of NPS with the high NPA of buccal and saliva samples, while saving cost, minimizing discomfort and reducing the risk of spread to the community and to HCWs.

**Figure 4.**
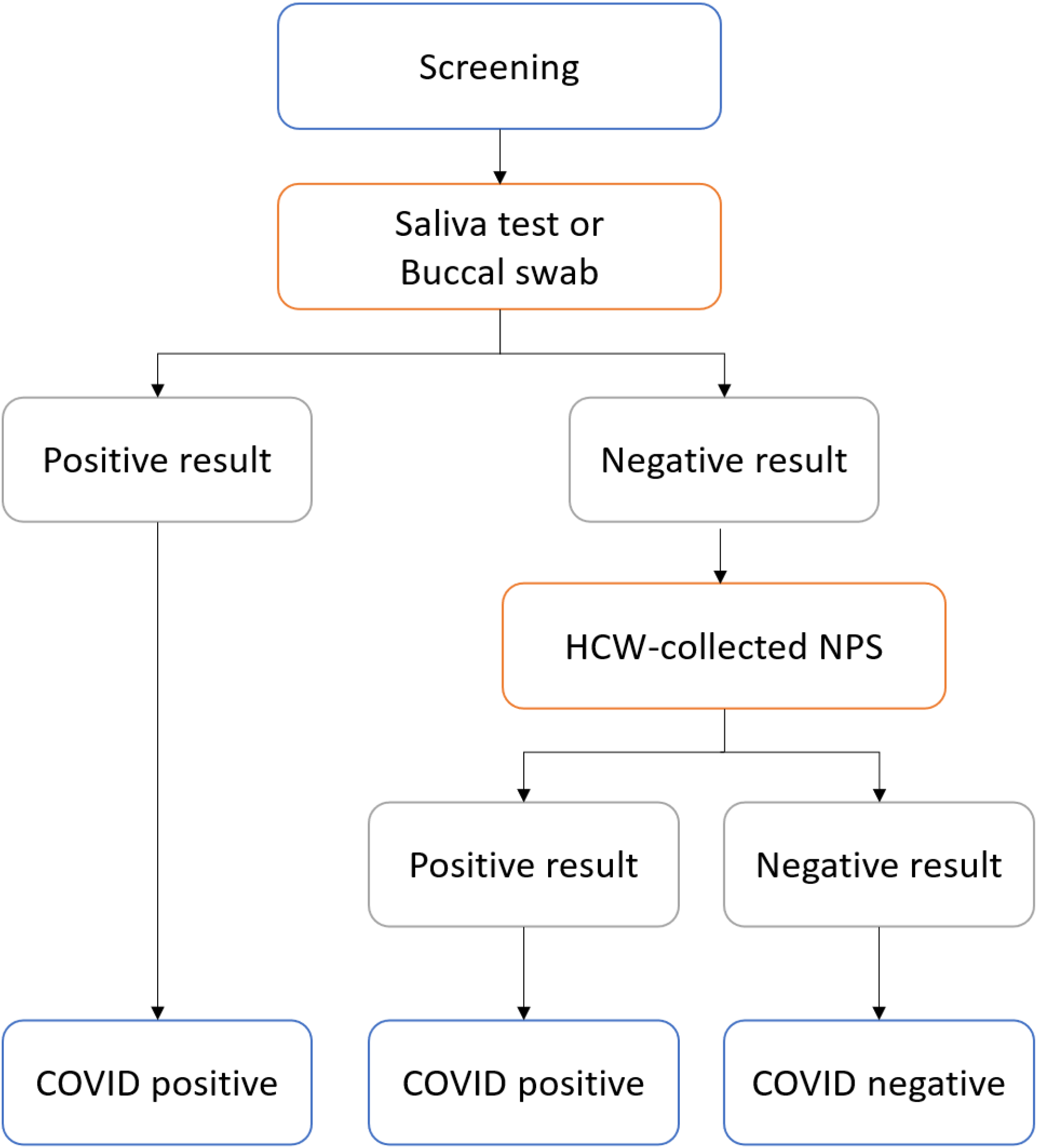
Recommended clinical workflow for population screening of SARS-CoV-2.

Our study has several strengths. Firstly, we were able to obtain self-collected specimens for buccal swab and saliva samples, testing the feasibility of widespread application in the community with saliva tests and buccal swabs being more acceptable to patients than NPS. Secondly, we were also able to recruit a variety of symptomatic and asymptomatic patients at various stages of their disease.

However, our study does have several limitations. Firstly, our sample size was relatively small. We have only included patients who previously tested positive for SARS-CoV-2, limiting its generalizability to population screening. Participants were also recruited up to a week after first testing positive for SARS-CoV-2 PCR, thus resulting in 26.2% who were negative on all three specimen samples. We also limited our study to the adult population. Chong et al has shown that saliva tests might not be useful in the paediatric population [25], which might limit its use as a population screening test. Even though the samples were self-collected, they were still conducted in the context of a healthcare setting and under the supervision of a HCW. Thus, the performance of the tests may be overestimated. Hence, larger validation studies need to be performed in both symptomatic and asymptomatic patients before buccal and saliva samples can be routinely recommended in the clinical setting.

## CONCLUSION

Saliva and buccal swabs were comparable to each other and were in moderate agreement with NPS for the detection of SARS-CoV-2. Both saliva and buccal swabs had greater patient preference in terms of comfort and convenience. Primary screening for SARS-CoV-2 may be performed with a saliva or buccal test. A negative test warrants a confirmatory NPS to avoid false negatives. Buccal swabs might be considered in the context of specific cohorts where spontaneous saliva production might be difficult. These self-collection methods represent feasible alternatives that could help reduce the discomfort experienced by patients, save costs, and reduce the risk of community spread and spread to HCWs. Larger trials should be conducted to determine the generalizability of these tests in both the symptomatic and asymptomatic population before they could be used for large scale community testing.

## Data Availability

The data that support the findings of this study are available on request from the corresponding author JCKY. The data are not publicly available due to them containing information that could compromise research participant privacy/consent.

## ACKNOWLEDGEMENTS

We would like to thank the patients for participating in the study. We would also like to thank the clinical research coordinators involved, Ms Ang Siew Boon and Mr Kearney Tan Jun Yao.

## Table

Table 1 Baseline demographics and clinical characteristics of patients and the results of paired nasopharyngeal, buccal and saliva test performed (n=42)

Table 2 Comparison between nasopharyngeal and buccal swab (a), and nasopharyngeal and saliva test

(b) with positive percent agreement, negative percent agreement, positive predictive value and negative predictive value.

Table 3 The associations between diagnostic tests and presence of symptoms

## FIGURE LEGENDS

Figure 2

- Nasopharyngeal swab
- Buccal swab
- Saliva test

